# Public reporting guidelines for outbreak data: Enabling accountability for effective outbreak response by developing standards for transparency and uniformity

**DOI:** 10.1101/2024.10.24.24315886

**Authors:** Vanessa Grégoire, Alex W. Zhu, Catherine M. Brown, John S. Brownstein, Denise Cardo, Fergus Cumming, Richard Danila, Christl A. Donnelly, Jeffrey S. Duchin, Mary-Margaret A. Fill, Christophe Fraser, Katie Fullerton, Sebastian Funk, Dylan George, Susan Hopkins, Moritz U.G. Kraemer, Marcelle Layton, Justin Lessler, Ruth Lynfield, James M. McCaw, Tristan D. McPherson, Zack Moore, Oliver Morgan, Steven Riley, Roni Rosenfeld, Erika Samoff, William Schaffner, Julie Shaffner, Roberta Sturm, Dawn Terashita, Henry Walke, Raynard E. Washington, Caitlin M. Rivers

**Affiliations:** Johns Hopkins Center for Health Security, Johns Hopkins University Bloomberg School of Public Health; Department of Environmental Health and Engineering, Johns Hopkins University Bloomberg School of Public Health; Massachusetts Department of Public Health; Harvard Medical School; Boston Children’s Hospital; Cardo MD Consultants, LLC; Division of Healthcare Quality Promotion, U.S. Centers for Disease Control and Prevention; UK Health Security Agency, and Foreign, Commonwealth and Development Office; Minnesota Department of Health; Department of Statistics and Pandemic Sciences Institute, University of Oxford; Public Health - Seattle & King County; Division of Infectious Diseases, University of Washington; School of Public Health; Division of Communicable and Environmental Diseases and Emergency Preparedness, Tennessee Department of Health; Pandemic Sciences Institute, University of Oxford; Surveillance and Data Modernization, Office of Readiness and Response, U.S. Centers for Disease Control and Prevention; London School of Hygiene & Tropical Medicine; Center for Forecasting and Outbreak Analytics, U.S. Centers for Disease Control and Prevention; UK Health Security Agency; Department of Biology, University of Oxford; Council of State and Territorial Epidemiologists; Department of Epidemiology, University of North Carolina Gillings School of Global Public Health; UNC Carolina Population Center; Department of Epidemiology, Johns Hopkins Bloomberg School of Public Health; School of Mathematics and Statistics, University of Melbourne; Centre for Epidemiology and Biostatistics, Melbourne School of Population and Global Health, University of Melbourne; Bureau of Communicable Diseases, New York City Department of Health and Mental Hygiene; Divisions of Infectious Diseases and Immunology, New York University Grossman School of Medicine; North Carolina Department of Health and Human Services, Division of Public Health; Pandemic and Epidemic Intelligence Systems, World Health Organization; Analytics and Surveillance, UK Health Security Agency; School of Public Health, Imperial College London; Machine Learning Department, School of Computer Science, Carnegie Mellon University; Vanderbilt University Medical Center; Career Epidemiology Field Officer assigned to the Tennessee Department of Health, U.S. Centers for Disease Control and Prevention; Communicable and Environmental Disease and Emergency Preparedness, Knox County Health Department; Acute Communicable Disease Control Program, Los Angeles County Department of Public Health; Office of Readiness and Response, U.S. Centers for Disease Control and Prevention; Mecklenburg County Public Health

**Keywords:** Epidemic, Outbreak, Data Reporting, Reporting Guidelines, Reporting Standards, Public Reporting

## Abstract

Currently, there are few standards for what essential information about an infectious disease outbreak should be reported to the public and when. The content and timeliness of public reporting (e.g. situation reports) is at the discretion of the jurisdiction overseeing the outbreak response, resulting in a substantial heterogeneity in available information. To address this problem, we undertook a consensus process to develop recommendations for what epidemiological information public health authorities should report to the public during an outbreak, including the administrative level and frequency of reporting. We first assembled a steering committee of nine experts representing federal public health, state public health, academia, and international partners to develop a candidate list of reporting items. We then invited 45 experts, 35 of whom agreed to participate in a Delphi panel. Of those, 25 participated in voting in the first round, 25 participated in voting in the second round, and 25 participated in voting in the third round, demonstrating consistent engagement in the consensus-building process. The final stage of the Delphi process consisted of a hybrid consensus meeting to finalize the voting items. This resulted in a final list of nine reporting items representing the minimum set of information to include in publicly available situation reports: Numbers of new confirmed cases, new hospital admissions, new deaths, cumulative confirmed cases, cumulative hospital admissions, and cumulative deaths, each reported weekly and at Administrative level 1 (typically state or province), and stratified by sex, age group, and race/ethnicity. This minimum reporting standard creates a strong framework and guidance for uniform sharing of outbreak information and promotes consistency of data between jurisdictions to enable prompt and effective response.

## Introduction

Situation reports are the regular updates that jurisdictions issue to describe the latest developments in infectious disease outbreaks. Situation reports, as well as other forms of public reporting, serve as vital sources of information for public health professionals in other jurisdictions, elected leaders, researchers, media, and the public.^1,2^ These reports often provide information that can guide the development and implementation of disease spread control measures.^1,2^ During the early days of the COVID-19 outbreak, for example, the information published by each jurisdiction was critical for helping other communities increase situational awareness and better understand the nature and trajectory of the growing pandemic.^3–5^

Currently, no guidelines exist to assist jurisdictions in deciding what and when to report.^6,7^ This has resulted in substantial heterogeneity in the information available across jurisdictions for various outbreaks. More uniform reporting would allow more consistent and reliable information to be available to stakeholders, enhancing the ability of decision makers to prepare for and respond to outbreaks.

To address this gap in guidance, we undertook a consensus process to develop recommendations for what epidemiological information public health authorities should report during an outbreak, including the administrative level and frequency of reporting. The Outbreak Reporting Best Practices in Transparency (ORBIT) guidelines aim to set a minimum standard for publicly available information during an outbreak to augment data reporting consistency and transparency. As part of this process, the research team also developed recommendations for types of outbreak situations where these guidelines would apply. However, we recognize the need for flexibility and professional judgment in these decisions.

As part of our efforts, we undertook a literature review to identify previous efforts to develop reporting guidelines. Although the research team identified several papers prioritizing epidemiological information, we did not find guidelines overlapping or conflicting with our project aim in the systematic review. To our knowledge, practitioners have only recently started developing standards for public reporting. The CORHA Principles and Practices for Healthcare

Outbreak Response, for example, provide guidance to public health and healthcare practitioners on when and what to notify patients and the public when an outbreak occurs in a healthcare setting, including how to communicate information in a way that helps mitigate risk.^8^ Additional information about the companion systematic review is available at https://doi.org/10.1101/2024.05.22.24307752.

## Methods

We followed the steps outlined in the Equator network’s guidance for the development of health research reporting guidelines.^9^ First, we assembled a steering committee of nine experts representing federal public health, state public health, academia, and international partners. Due to scheduling conflicts that precluded synchronous meetings, the principal investigator and secretariat met with steering committee members individually or in small groups to plan and finalize the project methodology.

The secretariat developed a preliminary list of candidate reporting items, which was informed by the literature review. Items proposed at this stage, for example, included confirmed, probable and suspected cases and deaths. We shared the draft list with steering committee members, who provided feedback regarding the relevance and wording of proposed items. Steering committee members also offered new candidate items for inclusion in the draft list.

After finalizing the preliminary list of candidate items, the research team assembled a Delphi^10^ panel to develop consensus on reporting items. This panel included 45 experts from various jurisdictions, expertise and career stages, who were invited via email to participate in a four-phase Delphi panel. Of the 45 invitees, 35 agreed to participate (Table 1 and Table 2). The secretariat sent each confirmed participant a spreadsheet with the candidate items. Participants were asked to assign each item on a scale of one to ten, with one being not important and ten being most important. Participants were instructed to vote only on the importance and practicality of each item, not the wording, and to remember that the list only represents the minimum set of reporting items. Of the 35 individuals that agreed to participate, 31 submitted a voting sheet and/or attended the consensus meeting.

**Table 1.** Participant jurisdictions. This table provides a broad overview of the countries and regions represented in the Delphi panel. The United States jurisdiction was further broken down into U.S. Federal and the respective Health and Human Services (HSS) Regions.

| **Jurisdictions** |  | **Number of individuals that agreed to participate** |
| --- | --- | --- |
| Australia |  | 1 |
| Hong Kong |  | 1 |
| South Africa |  | 1 |
| United States | HHS Region 1 | 2 |
|  | HHS Region 2 | 1 |
|  | HHS Region 3 | 1 |
|  | HHS Region 4 | 8 |
|  | HHS Region 5 | 2 |
|  | HHS Region 8 | 1 |
|  | HHS Region 9 | 1 |
|  | HHS Region 10 | 2 |
|  | U.S. Federal | 4 |
| United Kingdom |  | 7 |
| World Health Organization |  | 1 |
| Council of State and Territorial Epidemiologists |  | 2 |
| **Total** |  | **35** |

**Table 2.** Participant expertise. This table provides a broad overview of the range of expertise represented in the Delphi panel.

| **Expertise** | **Number of individuals that agreed to participate** |
| --- | --- |
| Administrative Level 0 (Federal) | 6 |
| Administrative Level 1 (typically state or province) | 9 |
| Administrative Level 2 (Local) | 6 |
| Academic | 11 |
| International | 1 |
| National | 2 |
| **Total** | **35** |

After a one-month voting period, the secretariat calculated the mean score for each item. Items that received a score of at least eight were judged to have reached consensus, and therefore admitted to the final stage of candidacy. Similarly, items that received a mean score less than or equal to five were dropped. Items with a score between five and eight were sent to participants for a second round of voting. This process was repeated for a total of three rounds of voting.

Of the expert participants who agreed to participate in the project, 25 participated in voting in the first round. Twenty-five participants returned round two spreadsheets, and 25 returned round three spreadsheets, demonstrating consistent engagement in the consensus-building process. Across all three rounds, 30 unique individuals participated in voting.

Following the three rounds of emailed voting, a final consensus meeting was held to finalize the reporting items. All Delphi participants were invited to attend the hybrid meeting in January 2024. Fifteen participants attended in person and seven participated online. The meeting aimed to discuss the remaining items that asynchronous voting had not resolved. The agenda also included time for participants to suggest changes to the wording of each item and to develop justifications for why each reporting item is important.

Meeting participants also discussed the types of outbreaks where these recommendations would be applicable. To resolve uncertainty, they engaged in a 45-minute discussion followed by a vote on the final wording of the recommendation. Items that received a score of at least eight were included in the final list. Therefore, the recommendations reflect the majority view, not the unanimous agreement of those present.

This study did not qualify as human subjects research according to the Johns Hopkins Bloomberg School of Public Health Institutional Review Board (IRB00023808).

## Recommendations

This process resulted in a final list of nine reporting items to include in publicly available situation reports:

1. The number of new confirmed cases registered during the previous reporting period. This item represents the number of new confirmed cases, as defined by the case definition (which should be published with the reporting items), reported to public health authorities since the last report. Although the diagnosis date is preferred, especially for epidemiological modeling and analytics, most jurisdictions will find that the reporting date is most feasible. Therefore, Delphi participants determine this to be an acceptable option and encourages readers to refer to Council of State and Territorial Epidemiologists (CSTE) guidance.^11^
2. The number of new hospital admissions related to the outbreak (among confirmed cases) registered during the previous reporting period. This item represents the number of new hospital admissions among confirmed cases, as defined by the case definition, reported to public health authorities since the last report.
3. The number of new deaths related to the outbreak (among confirmed cases) registered during the previous reporting period. This item represents the number of new deaths among confirmed cases, as defined by the case definition, reported to public health authorities since the last report.
4. The number of cumulative confirmed cases registered throughout each reporting period. This item represents the total number of confirmed cases, as defined by the case definition, reported to public health authorities since the start of the outbreak.
5. The number of cumulative hospital admissions related to the outbreak (among confirmed cases) registered throughout each reporting period. This item represents the total number of hospital admissions among confirmed cases, as defined by the case definition, reported to public health authorities since the start of the outbreak.
6. The number of cumulative deaths related to the outbreak (among confirmed cases) registered throughout each reporting period. This item represents the total number of deaths among confirmed cases, as defined by the case definition, reported to public health authorities since the start of the outbreak.
7. Establish weekly reporting. This item specifies that public health authorities should, at a minimum, report to the public weekly. This is a minimum recommendation; jurisdictions may exceed these standards at their discretion (e.g., report every day, every other day, etc.).
8. Establish Administrative level 1 (typically state or province) reporting of the final reporting items. This item specifies that public health authorities should, at minimum, report the data items to the public with state, province, or equivalent level geographic granularity.
9. Stratify reporting items by sex, age group, and race/ethnicity. This item specifies that public health authorities should stratify the reporting items by sex, age group, race/ethnicity, when doing so would not compromise privacy or confidentiality. Where and when the data is available, public health authorities should refer to their jurisdiction’s census bureau stratification for sex, age group, race/ethnicity.

### Deciding When to Apply the ORBIT Guidelines

Participants discussed and voted on the applicability of the ORBIT reporting guidelines. There was general agreement among participants that small, point-source outbreaks, such as an outbreak of foodborne illness in a restaurant, did not merit a public-facing situation report. On the other hand, participants expressed a reluctance to limit reporting to the most serious outbreaks, such as the novel coronavirus, in recognition that reliable reporting of more moderate outbreaks allows neighboring jurisdictions to take proactive preparedness and control measures based on early information.

Ultimately, participants reached consensus that an outbreak that necessitates reporting under the International Health Regulations (IHR) (2005) should trigger use of the reporting guidelines. Participants also recommend that for other outbreaks, the decision to trigger the reporting guidelines falls to in-country public health authorities.

Under Article 6 of the IHR (2005), State Parties must report events constituting potential public health emergencies of international concern (PHEIC) to the World Health Organization (WHO) if the situation of concern meets two of the following four criteria:^12,13^

1. Is the public health impact of the event serious?
2. Is the event unusual or unexpected?
3. Is there a significant risk of international spread?
4. Is there a significant risk of international travel or trade restrictions?

The WHO Director-General may then choose to declare a PHEIC based on advice from the IHR Emergency Committee, information provided by State Parties, scientific expert recommendations, and risks posed to human health, international disease spread and international travel.^12,14^

Participants agreed that this scenario adequately captures emerging outbreaks of high consequence. The definition also provides sufficient room for in-country public health authorities to exercise their judgment on emerging outbreaks that have not reached an international level of concern under the IHR (2005) but may evolve to outbreaks of higher consequence or still pose a significant risk of jurisdictional spread.

### Defining Confirmed Cases

Participants discussed the role of case definitions in the reporting guidelines extensively. They emphasized the need for nuance and flexibility in defining confirmed cases, depending on the outbreak stage. The ability to confirm cases often depends on the availability of laboratory tests. However, this may not be possible at the start of an outbreak with novel pathogens. For example, the first U.S. Centers for Disease Control and Prevention (CDC) diagnostic test kits for SARS-CoV-2 were approved by the U.S. Food and Drug Administration (FDA) on February 4, 2020, about two months after the WHO was notified of the first COVID-19 cases in China.^15,16^

Relying solely on laboratory confirmation assumes that tests are accessible and equitably distributed, which is often not the case. Unequal access to testing across U.S. cities and in low- and middle-income countries demonstrates this issue. Therefore, probable cases are important indicators to capture outbreak trends when confirmed diagnoses are difficult.^17–19^

To reflect this in the ORBIT guidelines, participants agreed that public health authorities should release information about probable or suspected cases at their discretion, considering outbreak timing, evolving case counts, and testing capacity. They also stressed the importance of coordination and transparency among federal, state and local authorities to ensure consistency across jurisdictions.

### Best Practices

These consensus guidelines provide public health jurisdictions with a minimum standard for what information to include in their regular reporting to the public. Public health jurisdictions may, however, choose to report additional data items at their discretion to further inform the public. Additional reporting recommendations discussed and generally supported amongst the participants include:

1. The median number of days from sample collection to the earliest known confirmed test result. This item represents an indicator for testing lag and has the potential to be subject to right truncation bias in an exponentially growing epidemic.^20^
2. Administrative level 2 (county or equivalent) reporting of the final reporting items. This item suggests that public health authorities should report the data items to the public with county or equivalent level geographic granularity when permissible considering the protection of privacy and preservation of confidentiality. As the consensus guidelines represent a minimum viable data set, jurisdictions may choose to exceed the reporting standards at their discretion by reporting county or equivalent level data alongside state, province, or equivalent data.
3. When the data is available and considerations of privacy and confidentiality permit it, public health authorities should report the onset date, followed by the specimen collection date and the diagnosis date, as per CSTE guidance, when reporting the number of new confirmed cases registered during the previous reporting period.^11^

## Public Health Implications

This set of minimum recommended reporting items creates a framework for uniform sharing of epidemic information and consistency of data between jurisdictions. The goal is to enable a more prompt and effective response by ensuring that the public, media, researchers, healthcare providers, and governments can access detailed information about an unfolding epidemic. As reporting needs may change over the course of an outbreak, we encourage coordination among public health authorities to maintain uniformity and comparability of reporting.

In future work, we will consider disease-specific iterations of these minimum reporting recommendations to increase reporting uniformity and usability. Additionally, we intend to align our current recommendations with other relevant efforts, such as those advanced by the U.S. CDC’s Data Modernization Initiative.^21^

Our guidelines were developed with input primarily from the United States and the United Kingdom, and participation from the WHO. We invite colleagues from other regions to develop similar processes that reflect their local contexts.

There are some limitations associated with the ORBIT guidelines. Participants contributing to the development process may have been influenced by their experiences with recent outbreaks, including COVID-19 and mpox, which may not fully reflect the needs of future outbreaks. Additionally, while collecting individual data, such as sex, age group, race/ethnicity, typically allows for more nuanced analysis than aggregate data, it also poses greater privacy and confidentiality risks. Race/ethnicity data, for instance, should be collected and communicated thoughtfully, with consideration for the protection of vulnerable groups. Lastly, the ORBIT guidelines remain a minimum standard. We encourage jurisdictions to consider reporting additional data that is most relevant and beneficial to the communities they serve.

In conclusion, these guidelines represent a minimum standard for what public health jurisdictions should include in their regular reporting to the public during an outbreak that meets IHR (2005) criteria or when in-country public health authorities have otherwise decided to trigger use of the guidelines. The standards detail minimum reporting items (e.g., the number of new confirmed cases), the frequency, and administrative level of reporting. We hope these standards facilitate timely, standardized reporting for critical public health events in recognition that such events pose a risk to the public and other public health jurisdictions who may wish to develop response plans.

## Data Availability

All data produced in the present study are available upon reasonable request to the authors

## Acknowledgements

The authors would like to acknowledge Erin Fink and Clint Haines for their contributions to data collection in the early stages of the project.

